# Association between physical activity and risk of incident cardiovascular disease in women with novel risk factors: evidence from the UK Biobank cohort

**DOI:** 10.1101/2024.10.14.24315492

**Authors:** Zhaleh Ataei, Leah Wright, Sergio Ruiz-Carmona, Erin J Howden

**Affiliations:** Baker Heart and Diabetes Institute, Melbourne, Australia; Baker Department of Cardiometabolic Health, The University of Melbourne; Centre for Health Analytics, Melbourne Children’s Campus, Melbourne, Australia

**Keywords:** pregnancy, menopause, pregnancy complications, primary prevention, exercise, cardiovascular disease

## Abstract

**Background:** Previous evidence demonstrates an increased risk of incident cardiovascular disease (CVD) in women with early menopause and/or adverse pregnancy outcomes. It also suggests an inverse association between physical activity and risk of incident CVD. However, this association in women with novel risk factors have not been evaluated. Therefore, we aimed to address this hypothesis.

**Methods:** We explored the data of 253,300 women without a history of CVD or life limiting conditions from the UK Biobank cohort, we further stratified women with a history of early menopause and/or complicated pregnancy to investigate how the risks affect indent CVD in women. We defined early menopause as any history of natural or surgical (bilateral oophorectomy) menopause < 47 years and adverse pregnancy outcomes as a history of gestational diabetes, hypertensive disorders of pregnancy, stillbirth or ≥ two miscarriages. We then classified women into three categories of physical activity based on their responses to the International Physical Activity Questionnaire. Women with a total MET-min/week of ≥ 3000, ≥ 600 and < 600 were categorised into high, moderate, and low levels of physical activity, respectively. We also assessed the dose-response interaction between physical activity and risk of incident CVD (heart failure (HF), arrhythmia, coronary heart disease (CHD), and stroke) in our population and adjusted all the models for potential CVD risk factors.

**Results:** During a median follow-up of 13 years, we found a significant increased risk of incident HF, arrhythmia and CHD in women with early menopause/complicated pregnancy, compared to women without these risk factors. Higher levels of physical activity reduced the risk of incident HF (HR 0.81, 95% CI 0.67 0.97) and arrhythmia (HR 0.84, 95% CI 0.75 0.94), compared to a low levels. Moreover, a moderate level of physical activity attenuated the risk of incident arrhythmia (HR 0.82, 95% CI 0.74 0.92). However, no significant associations were found between physical activity risk of incident stroke or CHD.

**Conclusion:** Our study indicates that higher levels of PA significantly attenuate the risk of HF and arrhythmia in women with novel risk factors and should be recommended to women to reduce their increased risk of CVD.

## INTRODUCTION

Cardiovascular disease (CVD) is the most common cause of mortality among women^1^. Despite recognition that gender and sex are important drivers of CVD risk, there remains an unmet need to understand what the female specific drivers of CVD outcomes and potential impacts of preventative approaches are. Some traditional risk factors, including diabetes, smoking and obesity, put women at higher risk of developing CVD than men^1–3^. The risk of developing coronary heart disease (CHD) conferred by obesity and cigarette smoking is greater in women compared to men by 18% and 25%, respectively^4^. Moreover, women with diabetes are at 44% higher risk of incident CVD compared to men.^2^ In addition, some unmodifiable or less modifiable stages in women’s lives, like early menopause and complicated pregnancy, make them even more vulnerable to CVD.^5,6^ According to previous studies, complicated pregnancy and early menopause affect up to 25%^7^ and ∼10%^8^ of women, respectively. However, currently there is poor recognition of these risk factors to overall CVD risk stratification and limited evidence of effective prevention approaches.

Physical activity is the cornerstone of CVD prevention. It reduces the risk of incident CVD and attenuates the harmful effects of other adverse behaviours, such as inappropriate sleep time or high sedentary behaviour, on cardiovascular health.^9–11^ Despite additional risk factors in women, they tend to be less active and are more sedentary than men (33.2% inactivity in women vs 29.9% in men)^2^. Physical activity is critical to preventing CVD in this large population. However, the effects of physical activity on the risk of incident CVD in women with novel risk factors, including early menopause and complicated pregnancy, has yet to be explored. Determining if physical activity is beneficial in this setting will have important public health implications, since engaging in physical activity is among the most simple and accessible methods to prevent CVD. Our aims were two-fold: firstly, to determine how female-specific risk factors influenced rates of CVD in women in the UK Biobank cohort. Secondly, to understand how physical activity modifies the increased risk of CVD in women with novel risk factors and to explore what the ideal level of physical activity is required to prevent developing CVD in this population. We hypothesised that physical activity would be a modifier of CVD outcomes in women with female-specific risk factors.

## METHODS

### Study design

The UK Biobank is a large prospective cohort study involving over 500,000 participants enrolled from 2006-2010 (Northwest Multicentre Research Ethics Committee (11/NW/0382), Resource number 55469)). At baseline, participants were aged 40-69 years old and assessed in various centres across England, Wales and Scotland and were followed up for different outcomes through linkages to national datasets. ^12^. Women with a previous history of CVD (N=8813), including heart failure (HF), coronary heart disease (CHD), arrhythmia, ischemic stroke, or any congenital heart disease were excluded from the study (Figure 1). We also excluded women with conditions that reduced their projected life expectancy to less than three years (Alzheimer’s disease, kidney failure, chronic obstructive pulmonary disease (COPD), and liver failure, N=3420). Information regarding the exact conditions and their related ICD-10 codes can be found in the supplementary section. The remaining 253,300 women were included in our study. Our analysis was divided into two stages to assess our aims and hypothesis: 1) To assess how novel risk factors impact the risk of developing CVD in women; and 2) To investigate how engaging in different levels of PA interacts with incident CVD in women with novel risk factors. For our second stage, we excluded women without a history of early menopause or complicated pregnancy in addition to those with missing physical activity data or responses that were considered as outliers according to guidelines^13^, giving us a final sample size of 40,952 women (Figure 1).

### Physical Activity Assessment

Physical activity was assessed according to the participants’ self-reported responses to the International Physical Activity Questionnaire (IPAQ)^14^. Individuals responded to questions regarding the amount and type of physical activity they engaged in the last seven days, including leisure time, domestic and gardening, as well as work and transport-related physical activity. A “high level” of physical activity was defined as achieving ≥ 3000 MET-min/week with a combination of different intensity activities or achieving ≥ 1500 MET-min/week with vigorous intensity activities. A moderate level of physical activity was defined as engaging in vigorous-intensity physical activity of ≥ 20 min/day for ≥ three days/week or doing a moderate-intensity activity for ≥ five days/week and/or walking of ≥ 20 min/day or achieving ≥ 600 MET-min/week by doing a combination of different intensity activities.

### Novel Risk factors

We defined novel risk factors as any history of early onset menopause (any natural or surgical (bilateral oophorectomy) menopause earlier than 47 years) and/or complicated pregnancy (positive history of one or more of the following: gestational diabetes, hypertensive disorders of pregnancy, stillbirth, and/or two or more miscarriages).^6,15^ Data regarding the number of miscarriages and stillbirths and the age of menopause were extracted from the self-reported questionnaires. Gestational diabetes was defined based on self-reported data and/or related ICD-10 codes. The data regarding hypertensive disorders of pregnancy were determined by ICD-10 codes.

### Endpoints

We defined the primary endpoints of this study as diagnosis of HF, CHD, arrhythmia, or ischemic stroke. All the events were reported using ICD-10 codes based on the data from hospitals/clinics (see table S1 supplementary material for complete list). If more than one event occurred to a participant, we only counted the first one. The number of events for each endpoint followed this rule, and only one event per participant was counted. Time to event for each participant was taken from the date of birth and censored at the date associated with the development of each endpoint or date of death or the date of their last follow-up session, whichever came first.

### Covariates

We adjusted the results for CVD risk factors, including age, smoking status, obesity, alcohol consumption, diabetes mellitus, hypertension, dyslipidaemia, and history of cancer. Data regarding the smoking status and alcohol intake frequency (how many times per day, week, or month) were obtained from self-reported questionnaires. Obesity was defined as BMI ≥ 30, calculated based on the participants’ responses to height and weight at their baseline visits. Diagnosis of hypertension, diabetes, dyslipidaemia, and cancer were defined by at least one of the related ICD-10 codes (supplementary section).

### Statistical analysis

We used Cox proportional hazard models to estimate the hazard ratios (HRs) and 95% confidence intervals (CIs) for incident CVD among women with and without novel risk factors. To determine how different levels of physical activity attenuate the risk of incident CVD we compared the effects of moderate and high levels of physical activity on the risk of incident CVD to the low-level physical activity in women with novel risk factors. Cox proportional hazard models were used to calculate the new hazard ratios and 95% confidence intervals. We evaluated the relationship between physical activity as a continuous variables and incident CVD with cubic spline models. We performed all statistical analyses with R version 4.2.1.

### Study ethics

The UK Biobank received ethical approval from the Northwest Multicentre Research Ethics Committee (11/NW/0382). All participants gave written informed consent. This study has been conducted using the UK Biobank Resource under application number 55469.

## RESULTS

### Baseline characteristic

Our study consisted of 253,300 women without a history of CVD or life limiting conditions from the UK Biobank cohort. Baseline characteristics of the included women are presented in table 1. The mean age at study entry was 57 ± 8 and 56 ± 8.0 years in women with and without novel risk factors, respectively. Overall, women without novel risk factors tended to be healthier, compared to women experiencing novel risk factors. Women with novel risk factors were more likely to smoke, and have obesity, diabetes, hypertension, dyslipidaemia, and increased incidence of past cancer. Of the 55,930 women with novel risk factors, 23,627 had a history of complicated pregnancy, and 35,838 experienced early menopause. There was overlap in novel risk factors in 3535 women. The median follow up was 13 years in which there were 8833 incidence CVD events in women with novel risk factors’ arm and 21804 in women with no novel risk factors (table 2). The incidence of CVD event rates were higher for each condition of interest among women with novel risk factors compared to those without these risk factors.

**Table 1.**
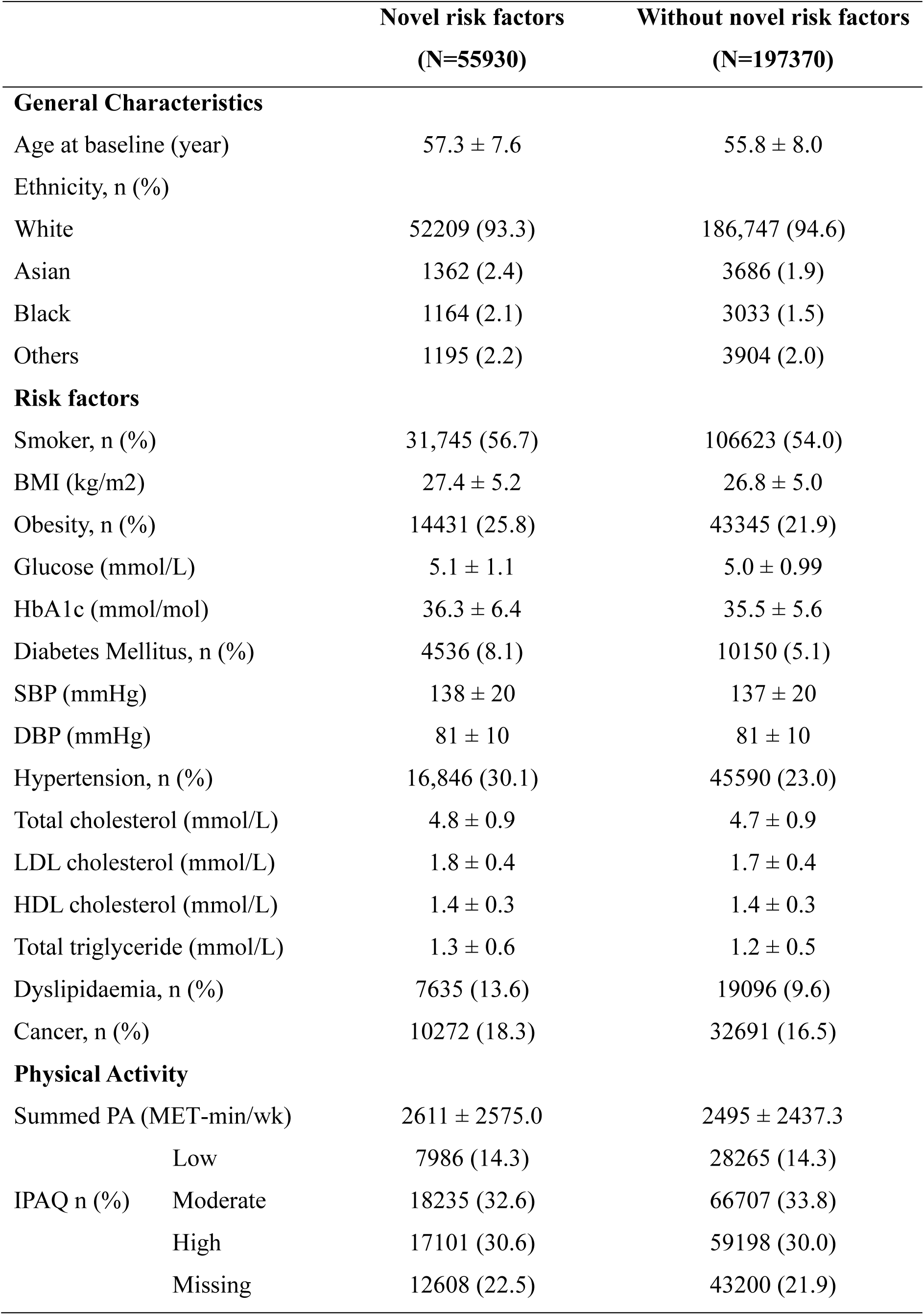
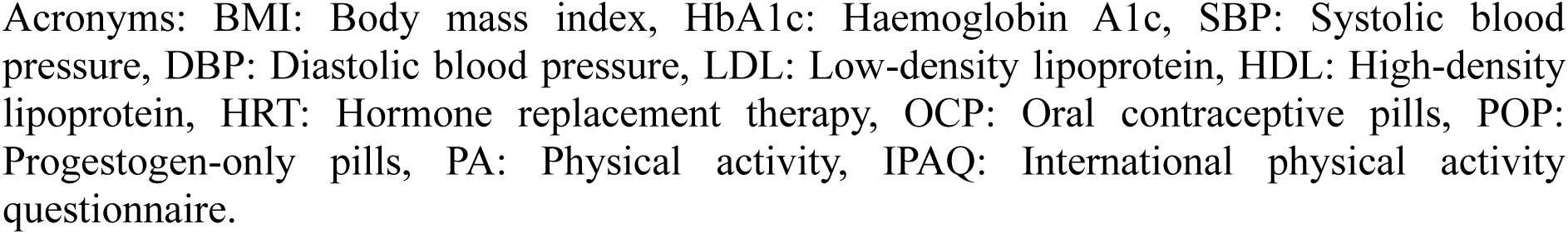
Participants Characteristics.

**Table 2.** Incidence of CVD Events in women with and without novel risk factors.

|  | **Novel risk factors** | **Without novel risk factors** |
| --- | --- | --- |
|  | **(N=55930)** | **(N=197370)** |
| Heart Failure, n (%) | 1374 (2.4) | 3318 (1.7) |
| Arrhythmia, n (%) | 2699 (4.8) | 7104 (3.5) |
| Coronary Heart Disease, n (%) | 3963 (6.6) | 9449 (4.7) |
| Ischemic stroke, n (%) | 797 (1.4) | 1933 (0.9) |

### Interaction between novel risk factors and CVD outcomes

In adjusted models, women with a history of early menopause had a significant increase in the risk of incident HF (HR 1.16, 95% CI 1.08 1.25), CHD (HR 1.15, 95% CI 1.10 1.20), arrhythmia (HR 1.09, 95% CI 1.04 1.14) and ischemic stroke (HR 1.18, 95% CI 1.07 1.29), compared to women without this risk factor. Similarly, women with a history of complicated pregnancy had a significantly higher risk of incident HF (HR 1.11, 95% CI 1.01 1.22), CHD (HR 1.17, 95% CI 1.11 1.23) and arrhythmia (HR 1.07, 95% CI 1.01 1.13), compared to ones without complicated pregnancy. However, there was no increased risk of ischemic stroke (HR 1.09, 95% CI 0.97 1.23) in the adjusted models.

### Physical activity and attenuation of heart failure risk in women with novel risk factors

In the unadjusted models, there was a significant association between physical activity levels and risk of incident HF demonstrating that both moderate (HR 0.69, 95% CI 0.59 0.82) and high (HR 0.59, 95% CI 0.49 0.70) levels of physical activity reduce the risk of incident HF significantly, compared to a low level of physical activity. However, after adjusting for confounders, only the association between a high level of physical activity and risk of developing HF remained significant (HR 0.81, 95% CI 0.68 0.97) (Figure 2). Dose-response models also showed a reduced risk of HF at a MET-min/week of 3000 (HR 1.10, 95% CI 0.94 1.28), compared to the baseline MET-min/week of 0 (HR 1.37, 95% CI 1.07 1.76) (Figure 6A).

### Physical activity and attenuation of arrhythmia risk in women with novel risk factors

In the unadjusted models, moderate (HR 0.69, 95% CI 0.62 0.77) and high (HR 0.67, 95% CI 0.60 0.75) levels of physical activity reduced the risk of arrhythmia, compared to low levels of physical activity. After adjusting for confounders, we observed a non-linear association in which the risk of arrhythmia decreased by 18% and 16% with moderate and high levels of physical activity, respectively (Figure 3). The dose-response association, demonstrated a lower risk of arrhythmia at MET-min/week of 3000 (HR 1.06, 95% CI 0.96 1.16) and 600 (HR 1.13, 95% CI 1.06 1.21), compared to 0 (HR 1.35, 95% CI 1.15 1.59) (Figure 6B).

### Physical activity and attenuation of coronary heart disease and stroke risk in women with novel risk factors

In the unadjusted models, higher levels of physical activity reduced the risk of incident CHD compared to a low level of physical activity; however, adjusting the models for confounders attenuated the effects (Figures 4). Similarly, the association between physical activity and risk of incident stroke was consistent with that observed for CHD and after adjusting for confounders, there was no statistically significant relationship between risk of incident stroke and physical activity (Figures 5).

### Impact of traditional risk factors on CVD outcomes in women with novel risk factors

In our multivariable models, traditional risk factors including hypertension and dyslipidaemia were significantly associated with increased risk across all cardiovascular outcomes. Hypertension demonstrated the strongest association with HF (HR 3.99, 95% CI 3.38 4.72), arrhythmia (HR 2.94, 95% CI 2.67 3.24) and stroke (HR 3.01, 95% CI 2.45 3.69) compared to other risk factors (Figures 2,3,5). While for CHD, dyslipidaemia (HR 3.90, 95% CI 3.58 4.24) emerged as the most significant determinant (Figure 4). Other factors, including diabetes, history of cancer, smoking and obesity were also included in the analysis; however, their associations with cardiovascular outcomes were less pronounced (Figure 2-5).

## DISCUSSION

The principal new findings in the new study are as follows: women with a history of novel CVD risk factors experience a higher frequency of CVD outcomes and have a higher burden of traditional CV risk factors than women without novel risk factors. Further in this cohort of higher risk women we found that higher levels of physical activity attenuated the risk of incident HF and arrythmia, but not CHD or ischemic stroke. These findings have important implications for public health recommendations for physical activity and highlight a high-risk group who could benefit from more targeted primary prevention strategies to increase physical activity levels in this population. Indeed, our analysis of the dose effect of physical activity on outcomes, identified that 3000 met minutes per week, which is consistent with current physical activity guidelines, reduced the risk of HF and arrythmia.

Both early menopause and complicated pregnancy are recognized contributors to an elevated risk of CVD. Although a multitude of female-specific factors have been labelled differently in many different studies, a consistent finding has been that female factors influence CVD outcomes, primarily through hormonal changes and cardiometabolic risk factors.^6,16^ The increased risk of CVD associated with early menopause is largely attributed to the premature loss of estrogen, a protective factor against CVD^16^. Consistent with these established mechanisms, our data showed that early menopause increased the risk of incident HF, CHD, arrhythmia, and stroke by 16%, 15%, 9%, and 18%, respectively. Similarly, pregnancy complications, including miscarriages ≥ 2, stillbirth, gestational diabetes, and hypertensive disorders of pregnancy, were associated with an increase in the risk of incident HF (11%), CHD (17%), and arrhythmia (7%). Women with a history of complicated pregnancy have an increased risk of developing CVD risk factors and later CVD^17^. The mechanism for this is likely multifactorial, with the development of CV risk factors mediated by a combination of lifestyle factors and vascular and metabolic dysfunction.

Accordingly, understanding the role of traditional risk factors is crucial for evaluating cardiovascular outcomes in this population. Hypertension is a strong CVD risk factor for premature mortality in women^18^ and women who have a history of hypertensive disorder of pregnancy have an increased risk of developing hypertension ^19^. In turn this relationship mediates the association with both HF and CAD in this patient group. Similarly, hyperlipidaemia is the driving factor of CHD development, without which the disease would not manifest.^20^ These patterns were evident in our study where hypertension emerged as the strongest contributor to an increased risk of HF, arrhythmia and ischemic stroke; while dyslipidaemia was the primary predictor of the risk of incident CHD, compared to other risk factors (Figure 2-5). These findings underscore the multifaceted nature of CVD and the importance of awareness of both traditional and novel risk factors to improve cardiovascular outcomes in this population. Emerging evidence suggest that optimal dosing of guideline directed medical therapy may differ for women^21^ and less women receive guideline directed therapy^22^. While we were unable to evaluate the usage of cardioprotective therapy in our study, our finding suggests a multi-dimensional approach is required to attenuate the increased risk of CVD.

This is the first study to our knowledge that has evaluated whether lifestyle factors influence CVD outcomes in women with novel risk factors. There is strong rationale for the promotion of physical activity to all women to mitigate CVD risk, but individuals with increased risk may benefit from more tailored intervention. Interestingly current physical activity guidelines are largely derived from studies of males^23^ and despite the growing understanding of sex and gender specific differences in responses to exercise^24,25^. there are no specific physical activity recommendations for healthy women, nor women with a high CVD risk. Therefore, studies that focus specifically on women and evaluate higher risk groups of women are essential to determine the relevance and effectiveness of current physical activity guideline and their effectiveness in preventing CVD. In this cohort we found that higher levels of physical activity lowered the risk of HF and arrhythmia incidence in women with novel risk factors. After adjusting for other confounders only the higher level of physical activity was protective for women developing HF. This is consistent with meta-analysis that reported that higher levels of physical activity are required to prevent HF ^26^. Conversely, in our adjusted models both moderate and higher levels of physical activity lowered the risk of arrhythmia, which is also consistent with prior findings demonstrating that more physically activity individuals have a lower risk of arrhythmia^27^ .

Previous studies have established the protective effects of physical activity on various cardiovascular outcomes. High levels of physical activity have consistently been associated with a significantly reduced risk of incident HF, with research indicating a 35% lower risk among individuals with the highest level of physical activity (four times guidelines recommended level), compared to a low levels.^28^ We demonstrated that a high level of physical activity (achieving ≥ 3000 MET-min/week through a combination of different intensities of physical activity) is protective against developing HF, compared to a low level of physical activity. On the other hand, our data showed both moderate (≥ 600 MET- min/week) and high levels of physical activity attenuate the risk of arrhythmia. Observing a non-linear relationship, our findings demonstrated that a moderate level of physical activity was the optimal level to prevent developing arrhythmia and no further significant risk reduction was achieved in women with a high level of physical activity, compared to those with a moderate level. This finding has also been observed elsewhere^29^. In contrast, a study from Azarbal et al., focusing on the data of nearly 82,000 postmenopausal women without a history of AF, observed a significant inverse association between a high level of recreational physical activity (> 9 MET-h/week) and risk of incident AF^30^. Variations in study population, domains of physical activity (recreational vs all domains) and different confounding factors might be the reason of having varied findings among the studies.

Interestingly, for CHD and stoke, no significant associations were observed after adjusting for potential confounding factors. While prior studies have observed that moderate ^31^ and high ^32^ ^33^ levels of physical activity attenuate the risk of incident CHD and stoke, our findings with a follow up of 13 years do not support this. It is possible that the discrepancy in findings with prior work is due to the shorter follow up in our study and difficulty in identifying patients with CHD or stoke prior to experiencing an event. For example, an arrhythmia can be detected through ECG screening, while an exercise or pharmacological stress test would be required to detect CHD and this is not routinely performed in HD and this is not routinely performed in *asymptomatic* people. Whereas, Hummel et al. found that higher levels of total physical activity(MET-h/day > 3.6) in women had a significantly lower risk of experiencing a myocardial infarction, compared to a low level of physical activity (MET-h/day ≤ 1.6)^32^. Apart from differences in study population, variations in the measurement of physical activity further complicate comparison between the studies.

The cardiovascular benefits of physical activity are largely due to the beneficial effects on CVD risk factors e.g. hypertension, dyslipidaemia, glycaemic control and optimal body composition, however, some of the mechanisms of benefits are yet to be determined ^34^. Sustaining higher levels of physical activity over a lifetime is likely to have provide critical cardioprotection for maintaining cardiovascular health women. Indeed, we have shown in women who initiate exercise training in early and mid-life, prior to menopause that exercise training reduces ventricular stiffness and improves cardiorespiratory fitness^25,35^ whereas other evidence suggests that cardiovascular benefits are blunted in women who increase physical activity levels after menopause^36^. Importantly women with a lifelong history of increased physical activity, consistent with the levels observed to be protective in this cohort of women with novel risk factors, had superior cardiac function and cardiorespiratory fitness than similar aged and younger sedentary women^37^. Physical activity can also contribute to lowering CVD risk, through better metabolic health with improved insulin sensitivity and glycaemic control ^38^, and better body composition^39^. The consequential impacts of which decreases the risk of increased adiposity which has been shown to directly increase inflammation and alter autonomic tone, thereby raising the risk of AF ^26,30,40^ and HF^41^.

### Strengths and limitations

To our knowledge, this is the first and largest study to assess the interaction between physical activity and risk of incident CVD in women with novel risk factors; however, some limitations must be acknowledged. The use of self-reported data may be subject to recall bias and there is a tendency to overestimate physical activity levels. We chose to evaluate total physical activity levels to reflect the overall weekly behaviour, recognising that for women in particular physical activity may be performed outside of structured settings. Physical activity behaviour was only assessed at baseline in our study, and therefore there may have been important changes to physical activity during follow-up that could have influenced our findings. Menopause history was also reported by self-report. Furthermore, while we adjusted our results for several key CVD risk factors, other risk factors, including female specific such as preterm delivery, intrauterine growth restriction and small for gestational age, or other risk factors including diet, sleeping habits and sedentary behaviour should be considered in future research. Further, the UK Biobank cohort may not be reflective of the general population, however there is greater than 50% participation from women which strengthens the generalisability for the focus of this study. Finally, in future studies it would be important to understand how novel risk factors contribute to different subtypes of HF e.g. heart failure with preserved ejection fraction which is 2-fold more common in women^42^ and what effects physical activity have in these populations.

## CONCLUSION

Our study highlights the effect of novel risk factors on CVD outcomes in women, while also presenting the protective effect of physical activity, specifically on incident HF and arrhythmia. We showed that guideline recommended physical activity levels of 150 minutes of moderate-vigorous physical activity or > 3000 met-minutes per week can prevent arrhythmia; however, to achieve the maximum protective benefit of physical activity against HF, women with novel risk factors need to engage in physical activity at twice the recommended levels. These findings reinforce the critical role of physical activity in mitigating cardiovascular risks and underscores the importance of educational campaigns designed for women with a history of novel risk factors to inform women about the potential benefits of physical activity and encourage them to be active. Moreover, our findings highlight the need for more precise guidelines regarding the optimal level of physical activity to effectively reduce the risk of incident CVD, specifically in this population.

## Data Availability

This research was conducted using the UK Biobank Resource (http://www.ukbiobank.ac.uk/) under application 55469. Data from the UK Biobank is accessible to eligible researchers via applying to www.ukbiobank.ac.uk.

## DECLARATIONS

### Author Contributions

Conception and design: EJH, LW, SRC

Collection and assembly of data: ZA

Data analysis and interpretation: ZA, EJH, LW, SRC

Manuscript writing: ZA (initial draft), All authors (revisions)

Final approval of manuscript: All authors

Accountable for all aspects of the work: All authors

## References

1. Vogel B, Acevedo M, Appelman Y, Bairey Merz CN, Chieffo A, Figtree GA, Guerrero M, Kunadian V, Lam CSP, Maas A, et al. The Lancet women and cardiovascular disease Commission: reducing the global burden by 2030. Lancet. 2021;397:2385–2438. doi: 10.1016/s0140-6736(21)00684-x

2. Garcia M, Mulvagh SL, Merz CN, Buring JE, Manson JE. Cardiovascular Disease in Women: Clinical Perspectives. Circ Res. 2016;118:1273–1293. doi: 10.1161/circresaha.116.307547

3. Sattar N, Rawshani A, Franzén S, Rawshani A, Svensson AM, Rosengren A, McGuire DK, Eliasson B, Gudbjörnsdottir S. Age at Diagnosis of Type 2 Diabetes Mellitus and Associations With Cardiovascular and Mortality Risks. Circulation. 2019;139:2228–2237. doi: 10.1161/circulationaha.118.037885

4. Huxley RR, Woodward M. Cigarette smoking as a risk factor for coronary heart disease in women compared with men: a systematic review and meta-analysis of prospective cohort studies. Lancet. 2011;378:1297–1305. doi: 10.1016/s0140-6736(11)60781-2

5. Roa-Díaz ZM, Raguindin PF, Bano A, Laine JE, Muka T, Glisic M. Menopause and cardiometabolic diseases: What we (don’t) know and why it matters. Maturitas. 2021;152:48–56. doi: 10.1016/j.maturitas.2021.06.013

6. Täufer Cederlöf E, Lundgren M, Lindahl B, Christersson C. Pregnancy Complications and Risk of Cardiovascular Disease Later in Life: A Nationwide Cohort Study. J Am Heart Assoc. 2022;11:e023079. doi: 10.1161/jaha.121.023079

7. Khan SS, Petito LC, Huang X, Harrington K, McNeil RB, Bello NA, Bairey Merz CN, Miller EC, Ravi R, Scifres C, et al. Body Mass Index, Adverse Pregnancy Outcomes, and Cardiovascular Disease Risk. Circ Res. 2023;133:725-735. doi: 10.1161/circresaha.123.322762

8. El Khoudary SR, Aggarwal B, Beckie TM, Hodis HN, Johnson AE, Langer RD, Limacher MC, Manson JE, Stefanick ML, Allison MA, et al. Menopause Transition and Cardiovascular Disease Risk: Implications for Timing of Early Prevention: A Scientific Statement From the American Heart Association. Circulation. 2020;142:e506–e532. doi: doi:10.1161/CIR.0000000000000912

9. Ekelund U, Steene-Johannessen J, Brown WJ, Fagerland MW, Owen N, Powell KE, Bauman A, Lee IM. Does physical activity attenuate, or even eliminate, the detrimental association of sitting time with mortality? A harmonised meta-analysis of data from more than 1 million men and women. Lancet. 2016;388:1302-1310. doi: 10.1016/s0140-6736(16)30370-1

10. Ekelund U, Brown WJ, Steene-Johannessen J, Fagerland MW, Owen N, Powell KE, Bauman AE, Lee IM. Do the associations of sedentary behaviour with cardiovascular disease mortality and cancer mortality differ by physical activity level? A systematic review and harmonised meta-analysis of data from 850 060 participants. Br J Sports Med. 2019;53:886-894. doi: 10.1136/bjsports-2017-098963

11. Chen LJ, Hamer M, Lai YJ, Huang BH, Ku PW, Stamatakis E. Can physical activity eliminate the mortality risk associated with poor sleep? A 15-year follow-up of 341,248 MJ Cohort participants. J Sport Health Sci. 2021. doi: 10.1016/j.jshs.2021.03.001

12. Sudlow C, Gallacher J, Allen N, Beral V, Burton P, Danesh J, Downey P, Elliott P, Green J, Landray M, et al. UK biobank: an open access resource for identifying the causes of a wide range of complex diseases of middle and old age. PLoS Med. 2015;12:e1001779. doi: 10.1371/journal.pmed.1001779

13. International Physical Activity Questionnaire. https://sites.google.com/view/ipaq/score.

14. Craig CL, Marshall AL, Sjöström M, Bauman AE, Booth ML, Ainsworth BE, Pratt M, Ekelund U, Yngve A, Sallis JF. International physical activity questionnaire: 12- country reliability and validity. Medicine & science in sports & exercise. 2003;35:1381–1395.

15. Zhu D, Chung HF, Dobson AJ, Pandeya N, Giles GG, Bruinsma F, Brunner EJ, Kuh D, Hardy R, Avis NE, et al. Age at natural menopause and risk of incident cardiovascular disease: a pooled analysis of individual patient data. Lancet Public Health. 2019;4:e553–e564. doi: 10.1016/s2468-2667(19)30155-0

16. Yoshida Y, Chen Z, Baudier RL, Krousel-Wood M, Anderson AH, Fonseca VA, Mauvais-Jarvis F. Early Menopause and Cardiovascular Disease Risk in Women With or Without Type 2 Diabetes: A Pooled Analysis of 9,374 Postmenopausal Women. Diabetes Care. 2021;44:2564–2572. doi: 10.2337/dc21-1107

17. Parikh NI, Gonzalez JM, Anderson CAM, Judd SE, Rexrode KM, Hlatky MA, Gunderson EP, Stuart JJ, Vaidya D, Epidemiology ObotAHACo, et al. Adverse Pregnancy Outcomes and Cardiovascular Disease Risk: Unique Opportunities for Cardiovascular Disease Prevention in Women: A Scientific Statement From the American Heart Association. Circulation. 2021;143:e902–e916. doi: doi:10.1161/CIR.0000000000000961

18. Gerdts E, Sudano I, Brouwers S, Borghi C, Bruno RM, Ceconi C, Cornelissen V, Diévart F, Ferrini M, Kahan T, et al. Sex differences in arterial hypertension: A scientific statement from the ESC Council on Hypertension, the European Association of Preventive Cardiology, Association of Cardiovascular Nursing and Allied Professions, the ESC Council for Cardiology Practice, and the ESC Working Group on Cardiovascular Pharmacotherapy. European Heart Journal. 2022;43:4777–4788. doi: 10.1093/eurheartj/ehac470

19. Honigberg MC, Zekavat SM, Aragam K, Klarin D, Bhatt DL, Scott NS, Peloso GM, Natarajan P. Long-Term Cardiovascular Risk in Women With Hypertension During Pregnancy. Journal of the American College of Cardiology. 2019;74:2743–2754. doi: doi:10.1016/j.jacc.2019.09.052

20. Sallam T, Watson KE. Predictors of cardiovascular risk in women. Womens Health (Lond). 2013;9:491–498. doi: 10.2217/whe.13.44

21. Agarwal A, Peters SAE, Chandramouli C, Lam CSP, Figtree GA, Arnott C. Guideline-Directed Medical Therapy in Females with Heart Failure with Reduced Ejection Fraction. Curr Heart Fail Rep. 2021;18:284–289. doi: 10.1007/s11897-021-00524-z

22. Sumarsono A, Xie L, Keshvani N, Zhang C, Patel L, Alonso WW, Thibodeau JT, Fonarow GC, Van Spall HGC, Messiah SE, et al. Sex Disparities in Longitudinal Use and Intensification of Guideline-Directed Medical Therapy Among Patients With Newly Diagnosed Heart Failure With Reduced Ejection Fraction. Circulation. 2024;149:510–520. doi: 10.1161/circulationaha.123.067489

23. Blair SN, LaMonte MJ, Nichaman MZ. The evolution of physical activity recommendations: how much is enough?1234. The American Journal of Clinical Nutrition. 2004;79:913S-920S. doi: 10.1093/ajcn/79.5.913S

24. Bassareo PP, Crisafulli A. Gender Differences in Hemodynamic Regulation and Cardiovascular Adaptations to Dynamic Exercise. Curr Cardiol Rev. 2020;16:65–72. doi: 10.2174/1573403x15666190321141856

25. Barnes JN, Fu Q. Sex-Specific Ventricular and Vascular Adaptations to Exercise. Adv Exp Med Biol. 2018;1065:329–346. doi: 10.1007/978-3-319-77932-4_21

26. Aune D, Schlesinger S, Leitzmann MF, Tonstad S, Norat T, Riboli E, Vatten LJ. Physical activity and the risk of heart failure: a systematic review and dose-response meta-analysis of prospective studies. Eur J Epidemiol. 2021;36:367–381. doi: 10.1007/s10654-020-00693-6

27. Elliott AD, Linz D, Mishima R, Kadhim K, Gallagher C, Middeldorp ME, Verdicchio CV, Hendriks JML, Lau DH, La Gerche A, et al. Association between physical activity and risk of incident arrhythmias in 402 406 individuals: evidence from the UK Biobank cohort. European Heart Journal. 2020;41:1479–1486. doi: 10.1093/eurheartj/ehz897

28. Pandey A, Garg S, Khunger M, Darden D, Ayers C, Kumbhani DJ, Mayo HG, de Lemos JA, Berry JD. Dose-Response Relationship Between Physical Activity and Risk of Heart Failure: A Meta-Analysis. Circulation. 2015;132:1786–1794. doi: 10.1161/circulationaha.115.015853

29. Jin MN, Yang PS, Song C, Yu HT, Kim TH, Uhm JS, Sung JH, Pak HN, Lee MH, Joung B. Physical Activity and Risk of Atrial Fibrillation: A Nationwide Cohort Study in General Population. Sci Rep. 2019;9:13270. doi: 10.1038/s41598-019-49686-w

30. Azarbal F, Stefanick ML, Salmoirago-Blotcher E, Manson JE, Albert CM, LaMonte MJ, Larson JC, Li W, Martin LW, Nassir R, et al. Obesity, physical activity, and their interaction in incident atrial fibrillation in postmenopausal women. J Am Heart Assoc. 2014;3. doi: 10.1161/jaha.114.001127

31. Armstrong ME, Green J, Reeves GK, Beral V, Cairns BJ. Frequent physical activity may not reduce vascular disease risk as much as moderate activity: large prospective study of women in the United Kingdom. Circulation. 2015;131:721–729. doi: 10.1161/circulationaha.114.010296

32. Hummel M, Hantikainen E, Adami HO, Ye W, Bellocco R, Bonn SE, Lagerros YT. Association between total and leisure time physical activity and risk of myocardial infarction and stroke - a Swedish cohort study. BMC Public Health. 2022;22:532. doi: 10.1186/s12889-022-12923-5

33. Sattelmair JR, Kurth T, Buring JE, Lee IM. Physical activity and risk of stroke in women. Stroke. 2010;41:1243–1250. doi: 10.1161/strokeaha.110.584300

34. Mora S, Cook N, Buring JE, Ridker PM, Lee IM. Physical activity and reduced risk of cardiovascular events: potential mediating mechanisms. Circulation. 2007;116:2110–2118. doi: 10.1161/circulationaha.107.729939

35. Howden EJ, Sarma S, Lawley JS, Opondo M, Cornwell W, Stoller D, Urey MA, Adams-Huet B, Levine BD. Reversing the Cardiac Effects of Sedentary Aging in Middle Age-A Randomized Controlled Trial: Implications For Heart Failure Prevention. Circulation. 2018;137:1549–1560. doi: 10.1161/circulationaha.117.030617

36. Seals DR, Nagy EE, Moreau KL. Aerobic exercise training and vascular function with ageing in healthy men and women. J Physiol. 2019;597:4901–4914. doi: 10.1113/jp277764

37. Carrick-Ranson G, Howden EJ, Brazile TL, Levine BD, Reading SA. Effects of aging and endurance exercise training on cardiorespiratory fitness and cardiac structure and function in healthy midlife and older women. Journal of Applied Physiology. 2023;135:1215–1235. doi: 10.1152/japplphysiol.00798.2022

38. Thyfault JP, Bergouignan A. Exercise and metabolic health: beyond skeletal muscle. Diabetologia. 2020;63:1464–1474. doi: 10.1007/s00125-020-05177-6

39. Westerterp KR. Exercise, energy balance and body composition. European Journal of Clinical Nutrition. 2018;72:1246–1250. doi: 10.1038/s41430-018-0180-4

40. Ofman P, Khawaja O, Rahilly-Tierney CR, Peralta A, Hoffmeister P, Reynolds MR, Gaziano JM, Djousse L. Regular physical activity and risk of atrial fibrillation: a systematic review and meta-analysis. Circ Arrhythm Electrophysiol. 2013;6:252–256. doi: 10.1161/circep.113.000147

41. Lopez-Jimenez F, Almahmeed W, Bays H, Cuevas A, Di Angelantonio E, le Roux CW, Sattar N, Sun MC, Wittert G, Pinto FJ, et al. Obesity and cardiovascular disease: mechanistic insights and management strategies. A joint position paper by the World Heart Federation and World Obesity Federation. European Journal of Preventive Cardiology. 2022;29:2218–2237. doi: 10.1093/eurjpc/zwac187

42. Gerber Y, Weston SA, Redfield MM, Chamberlain AM, Manemann SM, Jiang R, Killian JM, Roger VL. A Contemporary Appraisal of the Heart Failure Epidemic in Olmsted County, Minnesota, 2000 to 2010. JAMA Internal Medicine. 2015;175:996-1004. doi: 10.1001/jamainternmed.2015.0924

